# The machine learning algorithm identified COL7A1 as a diagnostic marker for LUSC and HNSC

**DOI:** 10.1101/2023.07.19.23292914

**Authors:** Chenyu Wang, Yongxin Ma, Jiaojiao Qi, Xianglai Jiang

## Abstract

Squamous cell carcinomas (SCCs) comes from different parts, but there may be similar tumorigenic signaling pathways and metabolism, and different squamous cell carcinoma has a similar mutation landscape and squamous differentiation expression. Studying the expression profile of common SCCs is helpful to find biomarkers with diagnostic and prognostic significance for a variety of squamous cell carcinoma. Lung squamous cell carcinoma (LUSC), head and neck squamous cell carcinoma (HNSC), and ‘squamous cell cancer’ in esophageal carcinoma (ESCA) and cervical squamous cell carcinoma and endocervical adenocarcinoma (CESC) in The Cancer Genome Atlas (TCGA) database were used as training sets. The relevant data sets in the Gene Expression Omnibus (GEO) database were selected as validation sets. Machine learning algorithms were used to screen out factors with high accuracy in the diagnosis of SCCs as core genes, and explore their effects on patient prognosis and immunotherapy. COL7A1 (Collagen Type VII Alpha 1 Chain) has high accuracy in the diagnosis of LUSC and HCSC, whether in the training set (LUSC _ AUC: 0.987; HNSC _ AUC: 0.933) or validation set (LUSC _ AUC: 1.000; HNSC _ AUC: 0.967). Moreover, the expression of COL7A1 was significantly correlated with shorter OS and DSS in HNSC and LUSC patients, and was also significantly negatively correlated with IPS in LUSC patients treated with CTLA4 (-) PD1 (+), CTLA4 (+) PD1 (-) and CTLA4 (+) PD1 (+). COL7A1 has the potential to be used as a diagnostic and prognostic marker for LUSC and HNSC and to predict the efficacy of LUSC immunotherapy.

## Introduction

Squamous cell carcinomas (SCCs) occur in epithelial cells and squamous cells. Common lesions include skin, esophagus, cervix, vagina, bronchus, etc^1-6^. The known causes of squamous cell carcinoma are diverse, including gene mutations, age, ultraviolet and human papillomavirus infection^3, 7^. SCCs with high incidence mainly include esophageal squamous cell carcinoma (ESCC), head and neck squamous cell carcinoma (HNSC), lung squamous cell carcinoma (LUSC) and cervical squamous cell carcinoma (CSCC). Lung squamous cell carcinoma is derived from bronchial epithelium and belongs to non-small cell lung cancer (NSCLC), and the 5-year survival rate of patients with LUSC is less than 20 %^8^. The diagnosis of LUSC and lung adenocarcinoma (LUAD) can be identified by detecting whether the immunohistochemical staining results of P63, P40 and CK5 / 6 are positive^9^. HNSC is the sixth most common cause of cancer death in the world. Due to the high recurrence and metastasis rate of advanced HNSC, the prognosis of patients with advanced HNSC is still poor even with various treatments such as surgery, radiotherapy and chemotherapy^10^. Esophageal squamous cell carcinoma ESCC is the main subtype of esophageal cancer. ESCC is highly heterogeneous, and the 5-year survival rate is about 20 %^11, 12^. CESC is the second most common gynecological cancer; there is still a lack of good biomarkers to detect early CESC, and CESC is often found in the advance stage^13^. SCCs come from different parts, but may have similar tumorigenic signaling pathways and metabolism, and different SCCs have similar mutation landscapes (such as TP53, SOX2 and TP63) and squamous differentiation expression^14-18^. Studying the expression profile of common SCCs is conducive to the discovery of biomarkers with diagnostic and prognostic significance for a variety of squamous cells, to detect SCCs early to improve the therapeutic effect of SCCs patients.

Bioinformatics technology provides a new method for the screening of tumor markers. The analysis of high-throughput sequencing data using bioinformatics technology is more conducive to finding genes with diagnostic value. The machine learning algorithm LASSO regression and SVM-RFE were used to identify the core genes with diagnostic potential. The Receiver Operating Characteristic Curve (ROC curve) was drawn and the area under the ROC curve (AUC) was calculated. The core genes with the most diagnostic value were selected as the diagnostic markers of squamous cell carcinoma, and the possible role of core genes in the development of specific cancers was analyzed^19-21^.

## Method

### Collect and process datasets

The RNA-seq and corresponding clinical data of LUSC, HNSC, and SCCs samples in ESCA and CESC in the TCGA database were used as training sets. Similarly, datasets from GEO (GSE9850, GSE19188, GSE23400, GSE30784) were collected and the sva R package was used to perform batch correction for four datasets^22-25^. The Limma R software package was used to calculate the differential genes between squamous cell carcinoma tissues and adjacent tissues. Differential genes with | logFC | > 2 and p value < 0.05 were selected as validation sets.

### Machine learning algorithms screen core genes

LASSO regression can use the constructed penalty function to calculate the optimal model and prevent the model from overfitting. The genes formed by the optimal model analyzed by lasso regression were used as the core genes for the diagnosis of squamous cell carcinoma. SVM-RFE is a machine learning algorithm based on Embedded method, which can complete the sorting of feature genes while screening useful feature genes^19-21^. The LASSO model was used to screen the core genes with diagnostic value for squamous cell carcinoma. On this basis, the SVM-RFE algorithm was used to calculate the final gene set with diagnostic value. The ROC R software package was used to draw the ROC curve of each gene expression in the core gene set and calculate the area under the ROC curve. Genes with high accuracy in both training set and validation set were selected as diagnostic markers for squamous cell carcinoma. The AUC of the expression of diagnostic markers in HNSC, LUSC, CSCC and ESCC was calculated respectively, and cancers with high diagnostic rates were selected to further analyze the possible biological functions of diagnostic markers in these cancers.

### Clinical features and prognosis analysis

The expression of core genes in squamous cell carcinoma was selected, and the correlation between the expression of core genes and the patient ‘s age, gender, stage, T staging, N staging, M staging was completed. The data of OS, DSS, DFI and PFI in patients with squamous cell carcinoma were collected and divided into two groups according to the expression of core genes. The correlation between the expression of SCCs core genes and OS, DSS, DFI and PFI was completed.

### Methylation analysis

Methylation is a kind of epigenetic phenomenon and the main form of epigenetics^26^. CpG islands are generated by promoter methylation to silence genes, thereby regulating gene expression. Methylation does not change DNA sequences, but can change traits and pass on to the next generation^27^. The promoter methylation data of core genes in UALCAN website were collected to study the effect of core gene methylation on OS in patients with squamous cell carcinoma.

### Enrichment analysis

Gene ontology (GO) analysis involves biological processes, molecular functions and cellular components. Kyoto Encyclopedia of Genes and Genomes (KEGG) analysis, as a widely used database, includes information on currently known signaling pathways. The ‘c5.go.v7.4.symbols.gmt’ file and the ‘c2.cp.kegg.v7.4.symbols.gmt’ file from the Molecular Signatures Database (MSigDB) were downloaded and used to perform GSEA-based GO enrichment analysis and KEGG enrichment analysis on differential genes in high and low expression samples of core genes.

### Analysis of tumor microenvironment and immune cell infiltration

The tumor microenvironment includes the structure, function and metabolism of the tissue where the tumor is located. It is also related to the internal environment of the tumor cells themselves. The tumor microenvironment regulates the occurrence, development and metastasis of the tumor. As a complex and dynamically regulated system, tumor microenvironment is composed of tumor cells, immune cells and supporting cells. The Estimate R software package was used to calculate the correlation between the expression of core genes and StromalScore, ImmuneScore and ESTIMATEScore in specific squamous cell carcinomas using wilcox test. The TMIER method was used to calculate the correlation between core gene expression and infiltration of B cell, T cell CD4, T cell CD8, Neutrophil, Macrophage and DC.

### Analysis of Immunotherapy

As an indicator of tumor immunogenicity, immunophenoscore (IPS) can predict the effect of immunotherapy in cancer patients^28^. Immunotherapy data were obtained from the TCIA database, and the correlation between the expression of core genes and IPS treated with CTLA4 (-) PD1 (-), CTLA4 (-) PD1 (+), CTLA4 (+) PD1 (-) and CTLA4 (+)

## Results

### Determination of core genes

The limma package was used to screen the differential genes (227 in total). The results of lasso regression analysis showed that a total of 37 genes were identified (Fig.1.A). The svm-rfe algorithm showed that a total of ten genes (SPP1, VEGFD, IL33, GPIHBP1, KIF18B, CRNN, HJURP, COL7A1, TMEM132A, CHI3L2) were identified as core genes (Fig.1.B). The AUC results of each gene in SCCs showed that TMEM132A (AUC: 0.944), KIF18B (AUC: 0.975), HJURP (AUC: 0.976) and COL7A1 (AUC: 0.929) had a higher diagnostic rate for squamous cell carcinoma in the training set (TCGA dataset) (Fig.2.A-J). The analysis results in the validation set (GEO dataset) showed that only COL7A1 (AUC: 0.907) had a high diagnostic rate for SCCs, so COL7A1 was selected as the core gene for the diagnosis of squamous cell carcinoma (Fig.3.A-D). The analysis of COL7A1 in different squamous cell carcinomas showed that the AUC of COL7A1 in CESC was 0.968 (Fig.4.A), the AUC in ESCA was 0.537 (Fig.4.B), the AUC in HNSC was 0.933 (Fig.4.C) and the AUC in LUSC was 0.987 (Fig.4.D) in the TCGA dataset. The AUC of COL7A1 in CESC was 0.648 (Fig.4.E), in ESCA was 0.951 (Fig.4.F), in HNSC was 0.967 (Fig.4.G) and in LUSC was 1.000 (Fig.4.H) in GEO dataset. COL7A1 was selected as a diagnostic biomarker for HNSC and LUSC for further analysis.

**Figure 1:**
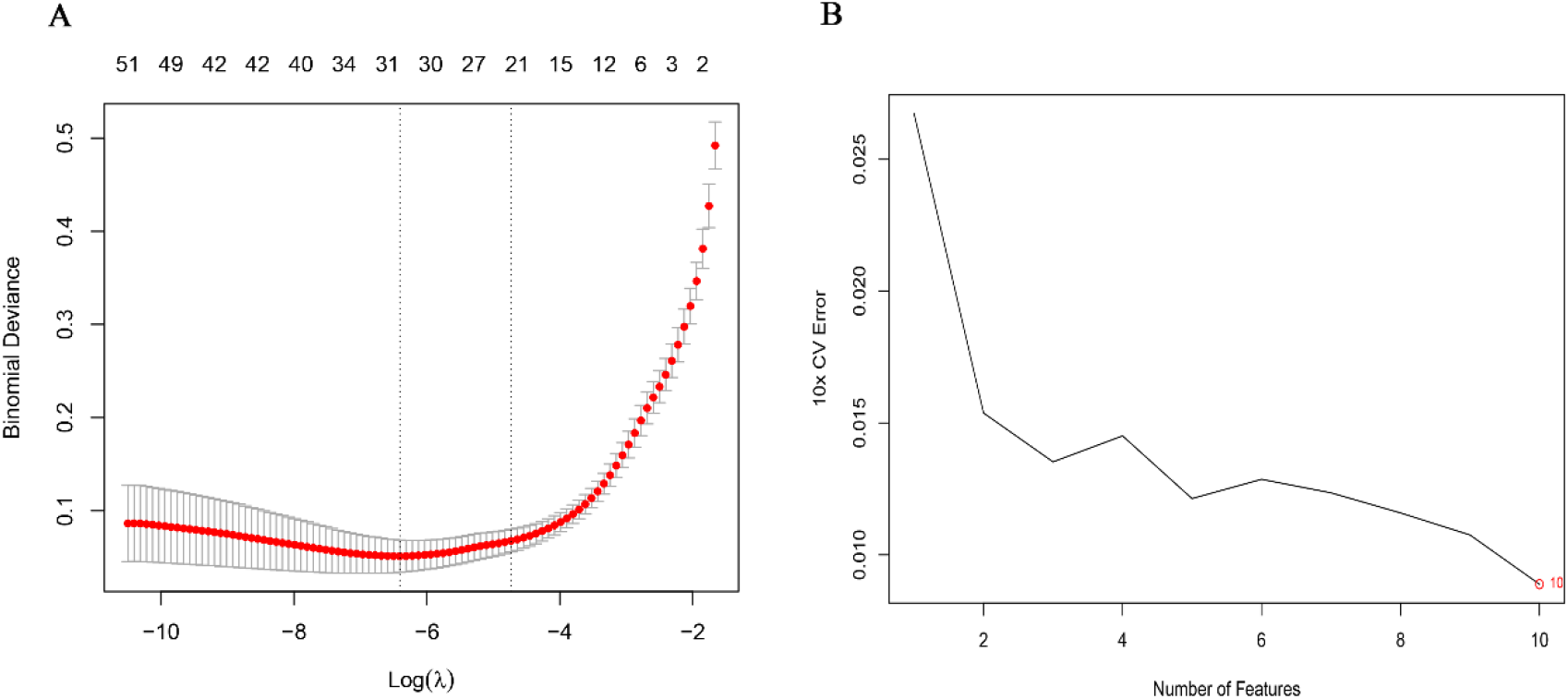
LASSO regression (A) and SVM-RFE (B) calculation results.

**Figure 2:**
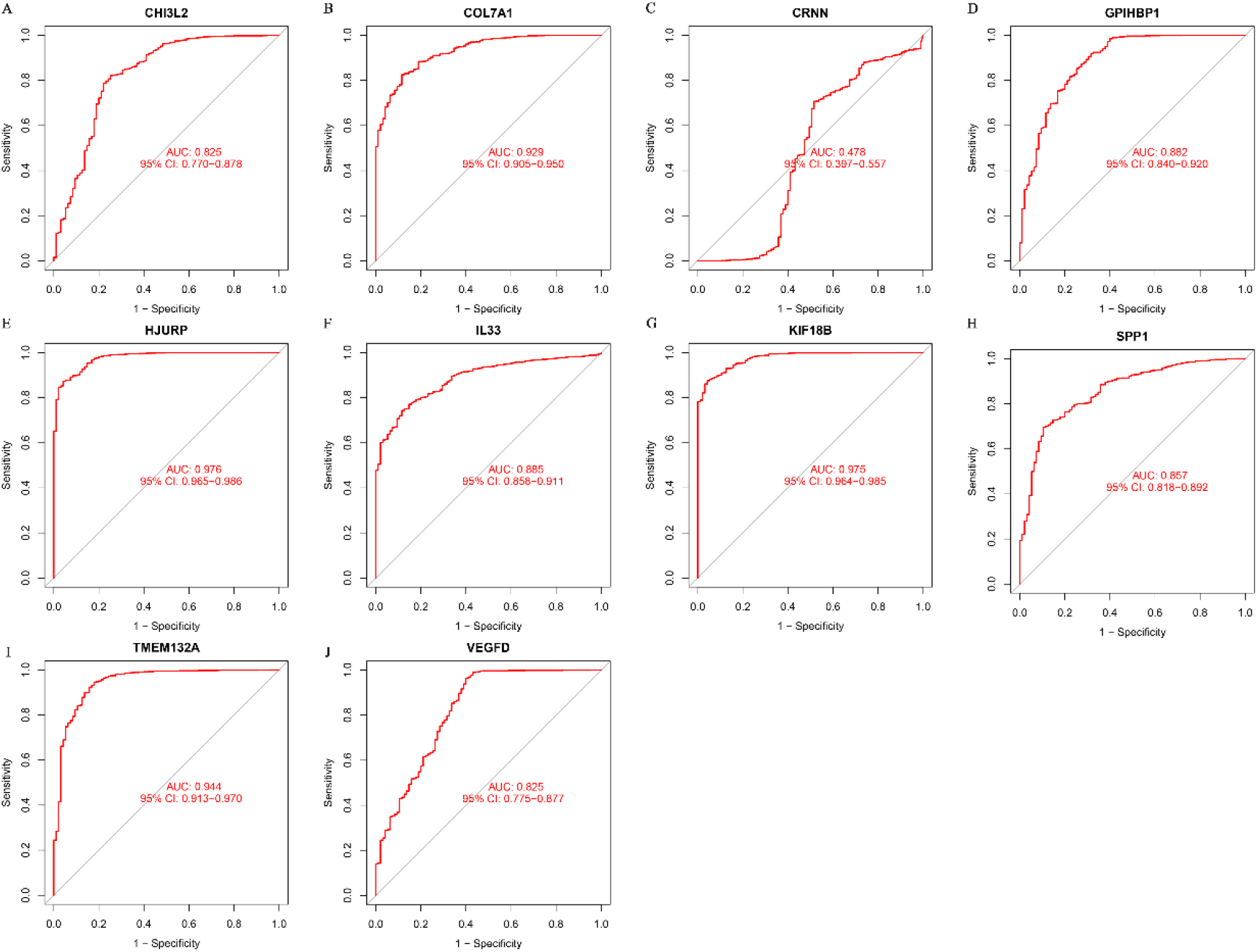
ROC curves of CHI3L2 (A), COL7A1 (B), CRNN (C), GPIHBP1 (D), HJURP (E), IL33 (F), KIF18B (G), SPP1 (H), TMEM132A (I) and VEGFD (J) in the diagnosis of SCCs in TCGA database.

**Figure 3:**
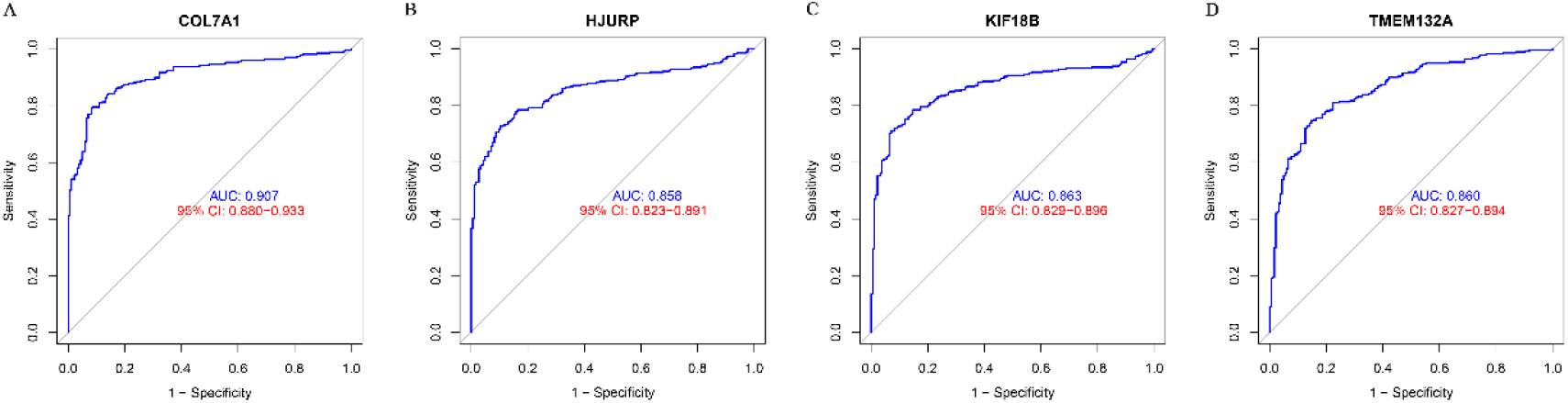
ROC curves of COL7A1 (A), HJURP (B), KIF18B (C) and TMEM132A (D) in the diagnosis of SCCs in GEO database.

**Figure 4:**
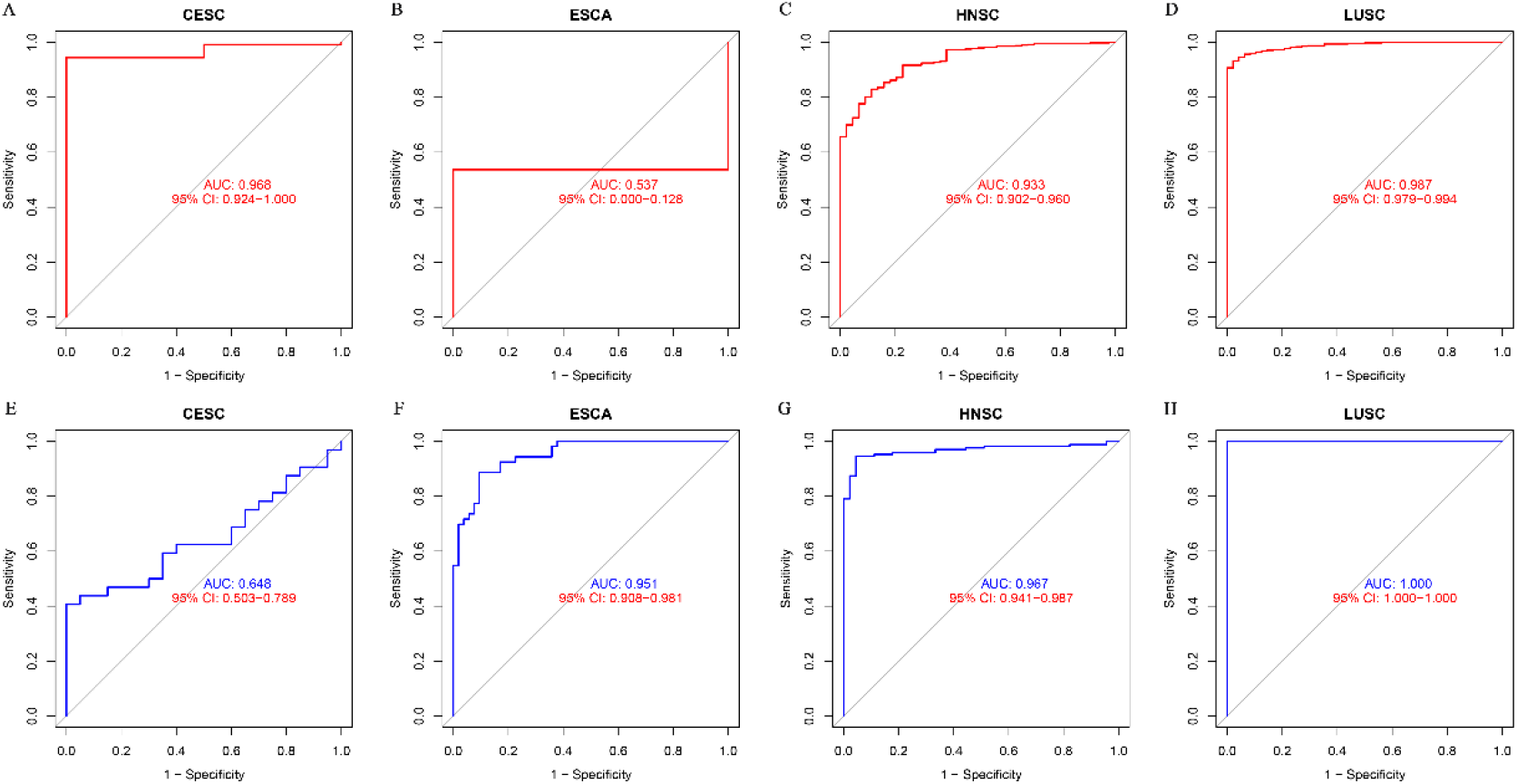
The ROC curves of COL7A1 in the diagnosis of CESC (A), ESCA (B), HNSC (C) and LUSC (D) in TCGA database; the ROC curve of COL7A1 in the diagnosis of CESC (E), ESCA (F), HNSC (G) and LUSC (H) in GEO database.

### Clinical features and prognosis analysis

The results of clinical characteristics analysis showed that there was no significant correlation between the expression of COL7A1 and age, gender, stage, T staging, N staging and M staging in LUSC patients and HNSC patients (Fig.5.A-F). The results of KM prognostic analysis showed that the high expression of COL7A1 was significantly positively correlated with shorter OS (Fig.6.A) and DSS (Fig.6.C) in LUSC patients and shorter OS (Fig.6.E) and DSS (Fig.6.G) in HNSC patients. There was no significant difference in DFI (Fig.6.B) and PFI (Fig.6.D) between LUSC patients with high and low expression of COL7A1, and there was no significant difference in DFI (Fig.6.F) and PFI (Fig.6.H) between HNSC patients with high and low expression of COL7A1.

**Figure 5:**
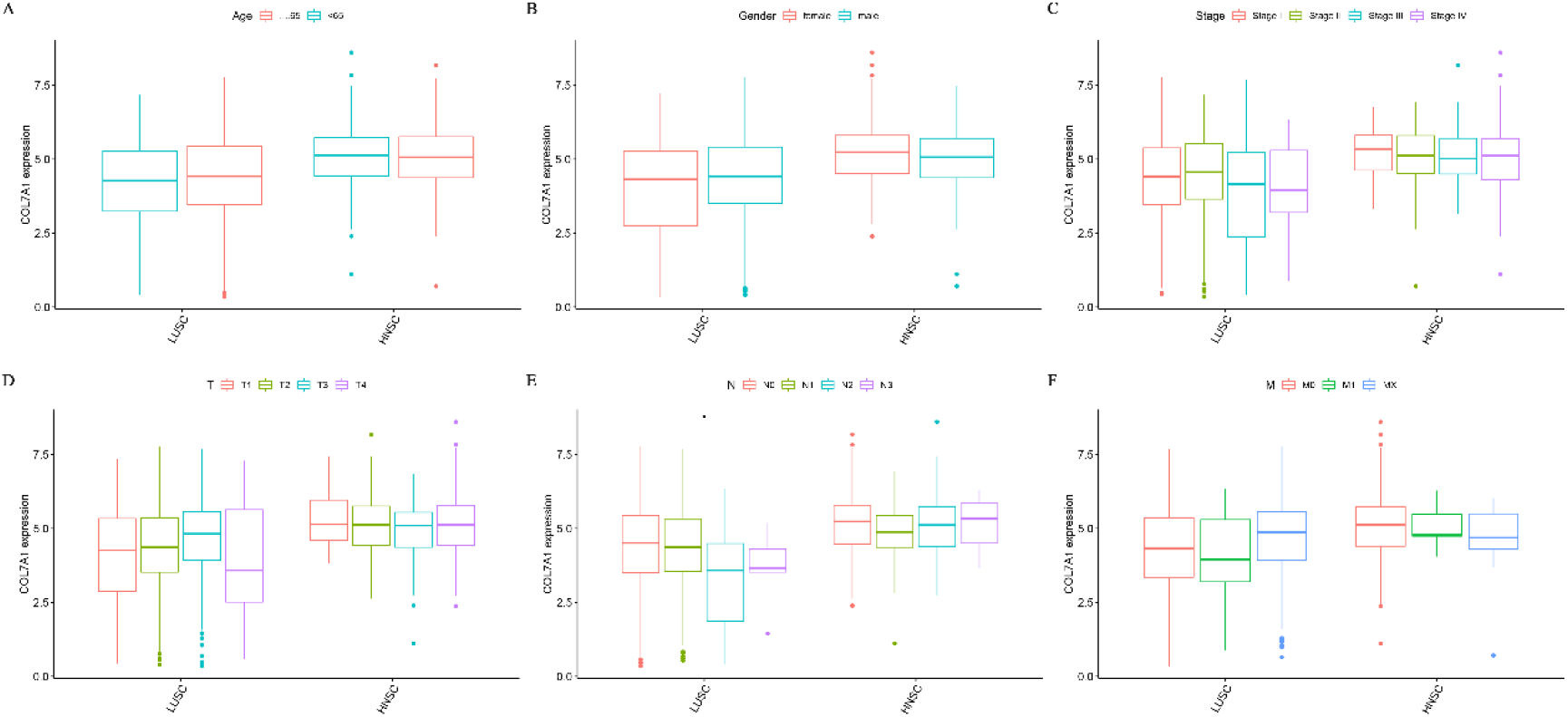
The correlation between COL7A1 expression and age (A), gender (B), stage (C), T stage (D), N stage (E) and M stage (F).

**Figure 6:**
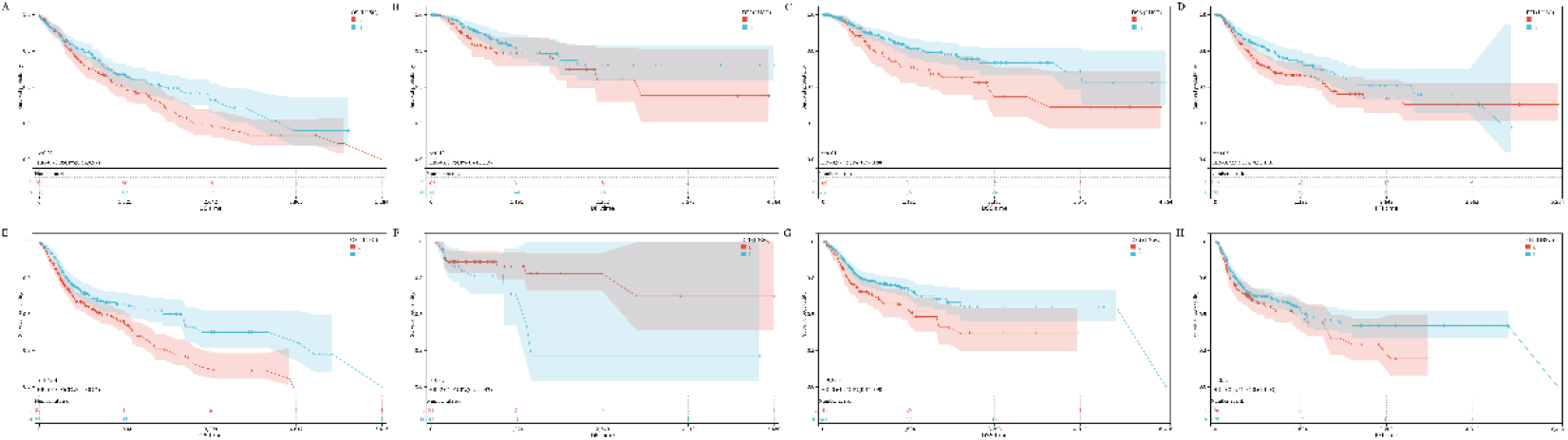
The correlation between the expression of COL7A1 and OS (A), DSS (B), DFI (C), PFI (D) in LUSC patients; the correlation between COL7A1 expression and OS (E), DSS (F), DFI (G), PFI (H) in HNSC patients.

### Promoter methylation level

The methylation level of COL7A1 in LUSC and HNSC obtained from UCSC xena website showed that COL7A1 had a lower Promoter methylation level in LUSC tumor (Fig.7.A-B).

**Figure 7:**
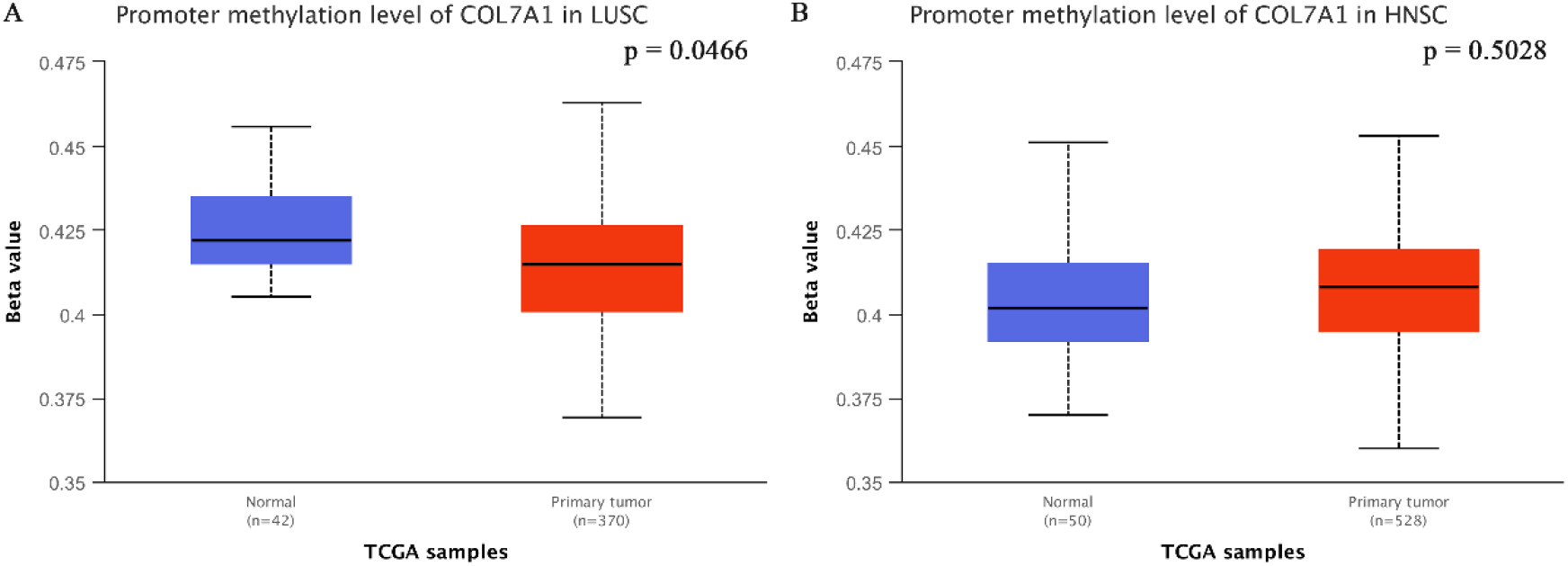
The difference of COL7A1 promoter methylation level between LUSC samples and adjacent samples (A); the difference of COL7A1 promoter methylation level between LUSC samples and adjacent samples (B).

### Enrichment analysis

Go enrichment analysis showed that the expression of COL7A1 was positively correlated with the activation of artery development, detection of chemical stimulus, sensory perception of smell, odorant binding and olfactory receptor activity in LUSC (Fig.8.A). And COL7A1 was positively correlated with the activation of etection of chemical stimulus, muscle cell proliferation, sensory perception of smell, transforming growth factor beta receptor signaling pathway and olfactory receptor activity in HNSC (Fig.8.C). KEGG enrichment analysis showed that the expression of COL7A1 was positively correlated with the activation of olfactory transduction, and negatively correlated with the activation of folate biosynthesis in LUSC (Fig.8.B). The expression of COL7A1 was positively correlated with the activation of olfactory transduction, and negatively correlated with the activation of O glycan biosynthesis, riboflavin metabolism and starch and sucrose metabolism in HNSC (Fig.8.D).

**Figure 8:**
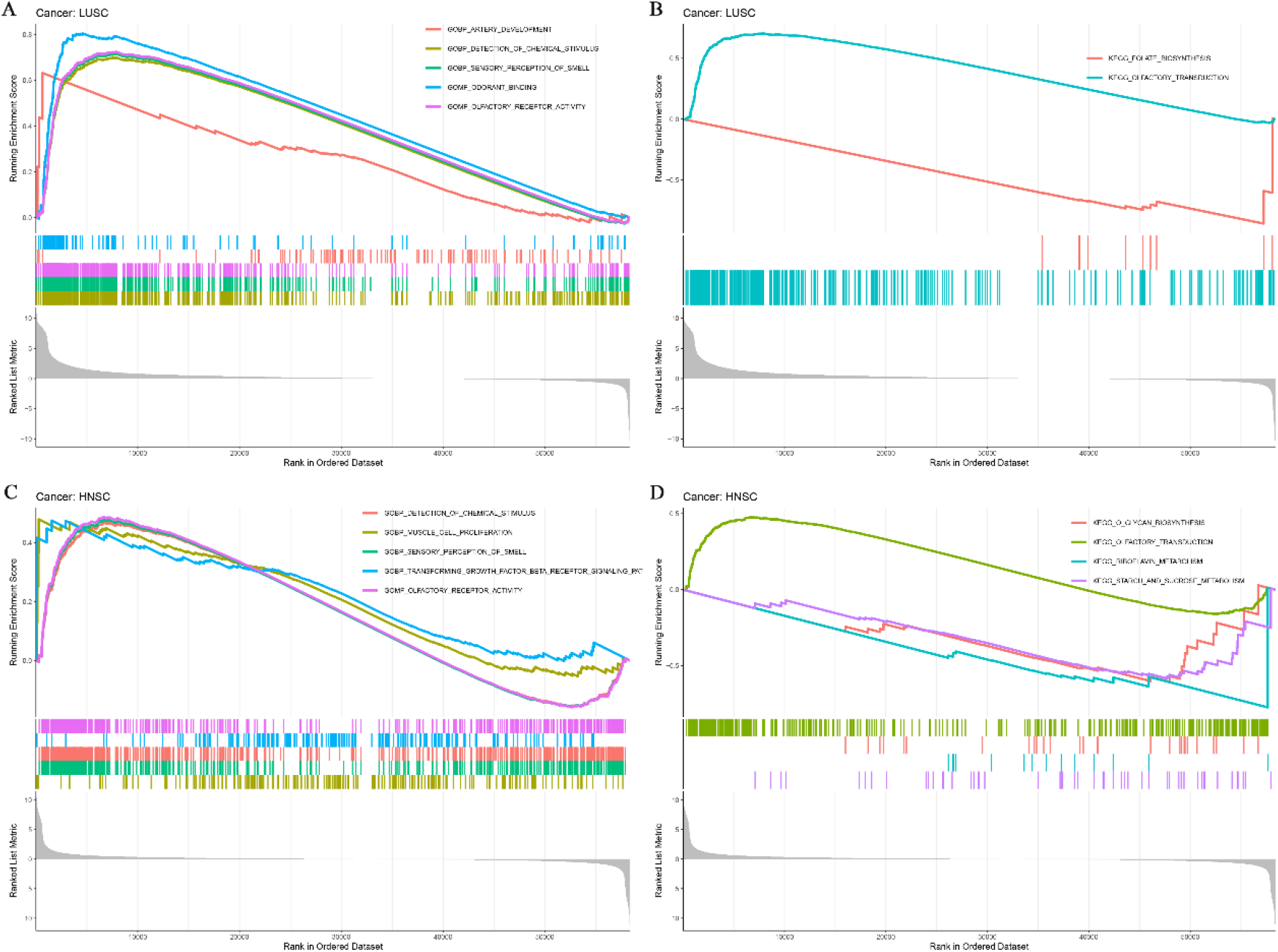
GO enrichment analysis (A) and KEGG enrichment (B) of COL7A1 in LUSC based on GESA; GO enrichment analysis (C) and KEGG enrichment (D) of COL7A1 in HNSC based on GESA.

### Tumor microenvironment and immune cell infiltration

The results of tumor microenvironment analysis showed that the StromalScore, ImmuneScore and ESTIMATEScore of LUSC samples in the low expression group of COL7A1 were significantly higher than those in the high expression group of COL7A1 (Fig.9.A). There was no significant difference in StromalScore, ImmuneScore and ESTIMATEScore between HNSC samples of COL7A1 low expression group and HNSC samples of COL7A1 high expression group (Fig.9.B). The results of immune cell infiltration analysis showed that the expression of COL7A1 was significantly negatively correlated with the infiltration of B cell, T cell CD8, Macrophage and DC in LUSC, and the expression of COL7A1 was significantly negatively correlated with the infiltration of B cell, T cell CD4, Macrophage and DC in HNSC (Fig.9.C).

**Figure 9:**
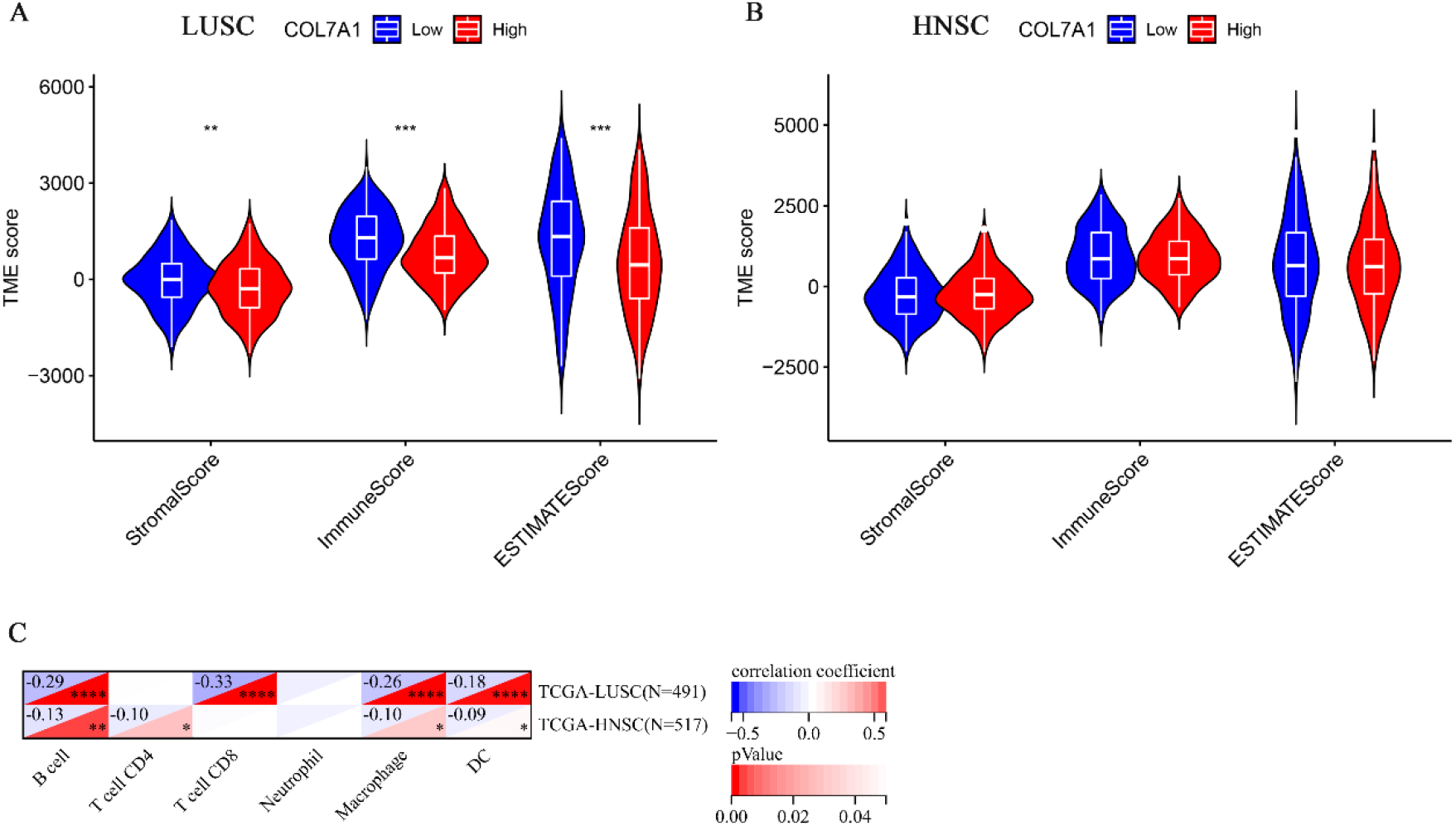
The correlation between the expression of COL7A1 and the tumor microenvironment score in LUSC (A) ; the correlation between the expression of COL7A1 and the tumor microenvironment score in HNSC (B) ; the correlation between COL7A1 expression and immune cell infiltration (C).

### Immunotherapy

Immunotherapy analysis from the TCIA database showed that the expression of COL7A1 was not significantly correlated with IPS treated with CTLA4 (-) PD1 (-) (Fig.10.A), and was significantly negatively correlated with IPS treated with CTLA4 (-) PD1 (+), CTLA4 (+) PD1 (-) and CTLA4 (+) PD1 (+) in LUSC (Fig.10.B-D). There was no significant correlation between IPS receiving CTLA4 (-) PD1 (-), CTLA4 (-) PD1 (+), CTLA4 (+) PD1 (-) and CTLA4 (+) PD1 (+) treatment in HNSC (Fig.10.E-H).

**Figure 10:**
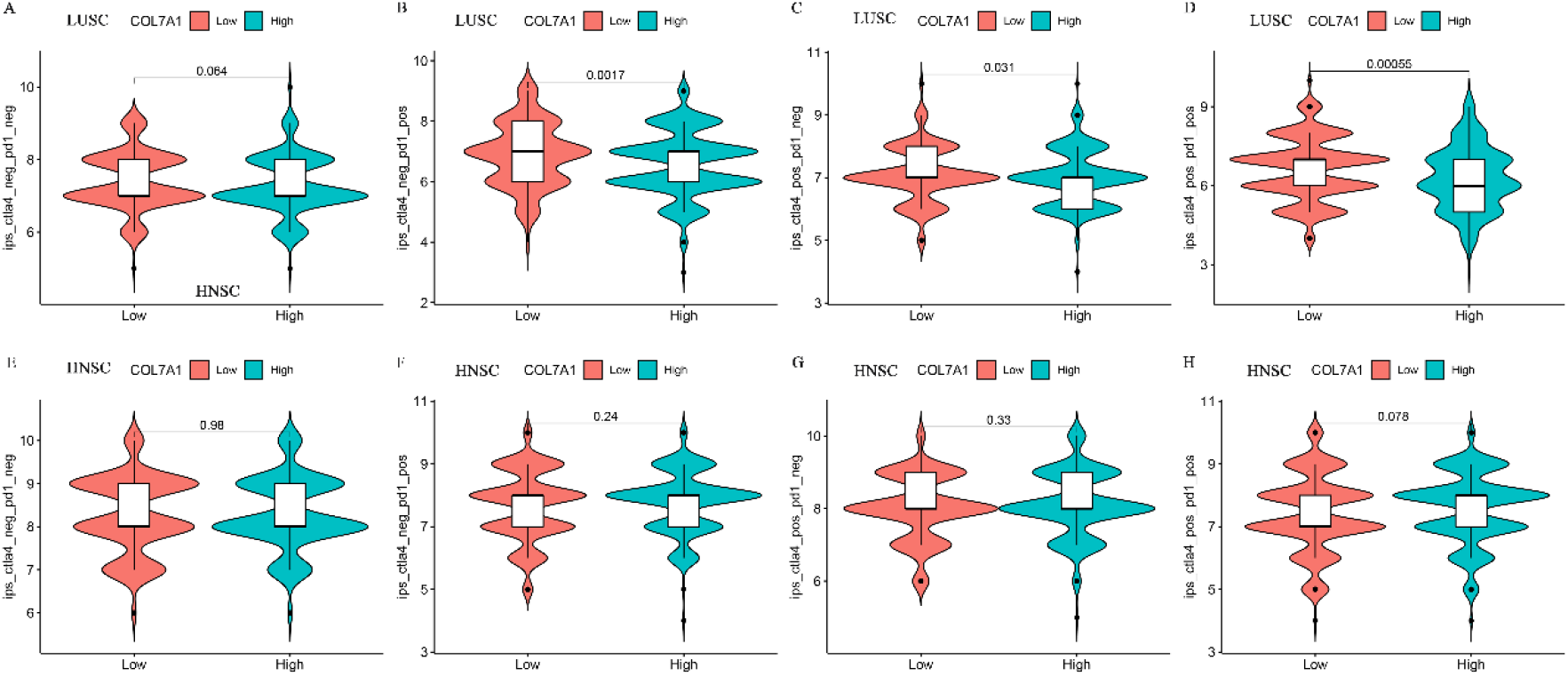
The correlation between COL7A1 expression and IPS in LUSC patients treated with CTLA4 (-) PD1 (-), CTLA4 (-) PD1 (+), CTLA4 (+) PD1 (-) and CTLA4 (+) PD1 (+) (A-D) ; the correlation between COL7A1 expression and IPS in HNSC patients receiving CTLA4 (-) PD1 (-), CTLA4 (-) PD1 (+), CTLA4 (+) PD1 (-) and CTLA4 (+) PD1 (+) treatment (E-H).

## Discussion

Even though multiple genes (COL7A1, HJURP, KIF18B and TMEM132A) with high accuracy in the training set were calculated by LASSO regression and SVM-RFE algorithm, only COL7A1 showed high accuracy in the validation set (AUC: 0.907). In specific cancer types, COL7A1 has poor prediction performance in ESCA samples in the training set and CESC samples in the validation set. However, COL7A1 has high accuracy in the diagnosis of HNSC and LUSC. Whether in the training set or the validation set, COL7A1 has the potential to be used as a diagnostic marker for HNSC and LUSC. The results showed that COL7A1 was worth exploring as a diagnostic marker for LUSC (AUC in the training set was 0.987 and AUC in the validation set was 1). COL7A1 encodes collagen VII (C1), which is involved in the assembly of anchoring fibrils that fix the epidermis and dermis^29^. Previous studies have shown that alteration of COL7A1 can cause Recessive dystrophic epidermolysis bullosa (RDEB), and RDEB can cause skin fragility and persistent blisters, and can rapidly progress to fibrosis and even SCCs^29, 30^. Compared with normal esophageal tissue, COL7A1 is highly expressed in ESCA.COL7A1 is significantly correlated with depth of tumor invasion and lymphatic invasion in ESCA, and the expression of COL7A1 is significantly negatively correlated with the 0-year survival rate of ESCA patients^31^. Although there was no significant correlation between the expression of COL7A1 and the age, gender and stage of patients including LUSC and HNSC patients, the results of prognostic analysis showed that the high expression of COL7A1 was significantly associated with shorter OS and DSS in HNSC and LUSC patients, indicating that COL7A1 also has the potential as a prognostic marker. In LUSC, the expression of COL7A1 was significantly negatively correlated with StromalScore, ImmuneScore and ESTIMATEScore, as well as the infiltration of B cell, T cell CD8, Macrophage and DC, indicating that COL7A1 affects the tumor microenvironment and tumor immunity of LUSC. Further immunotherapy analysis showed that the expression of COL7A1 was significantly negatively correlated with IPS in LUSC patients treated with CTLA4 (-) PD1 (+), CTLA4 (+) PD1 (-) and CTLA4 (+) PD1 (+), indicating that COL7A1 may also have the prospect of predicting the efficacy of LUSC immunotherapy.

Due to the unpredictability of the results of machine learning algorithms, our study has not yet obtained appropriate clinical tissues to verify its predictive performance as a diagnostic marker. However, our study includes different databases and data sets. The results of statistical calculations on the differential expression of COL7A1 in these cancer tissues and adjacent tissues are surprising, which provides a promising marker for the early diagnosis of LUSC and HNSC.

## Conclusion

COL7A1 has the potential to be a diagnostic marker for LUSC and HNSC (especially in LUSC). The high expression of COL7A1 is significantly correlated with shorter OS in HNSC patients and LUSC patients and is significantly negatively correlated with IPS in LUSC patients receiving CTLA4 (-) PD1 (+), CTLA4 (+) PD1 (-) and CTLA4 (+) PD1 (+) immunotherapy.

## Data Availability

The datasets analyzed during the current study are available in TCGA (https://portal.gdc.cancer.gov/), GEO database (https://www.ncbi.nlm.nih.gov/geo/), TISIDB database (http://cis.hku.hk/TISIDB/) and TCIA database (https://tcia.at/home).

https://www.ncbi.nlm.nih.gov/geo/

http://cis.hku.hk/TISIDB/

https://tcia.at/home

## Declarations

### Ethics approval and consent to participate

Not applicable

### Consent for publication

Not applicable

### Author Contributions

Xianglai Jiang and Chenyu Wang conceived the study, Yongxin Ma comprehensively collected relevant data, Chenyu Wang completed the work on data analysis, Xianglai Jiang completed the draft, Jiaojiao Qi and Xianglai Jiang reviewed the paper.

## Acknowledgement

This work has benefited from the aforementioned databases.

